# PFHpA alters lipid metabolism and increases the risk of metabolic dysfunction-associated steatotic liver disease in youth—a translational research framework

**DOI:** 10.1101/2024.07.01.24309775

**Authors:** Brittney O. Baumert, Ana C. Maretti-Mira, Zhenjiang Li, Nikos Stratakis, Yinqi Zhao, Douglas I. Walker, Hongxu Wang, Fabian Christoph Fischer, Qiran Jia, Damaskini Valvi, Scott M. Bartell, Carmen Chen, Thomas Inge, Justin Ryder, Todd Jenkins, Stephanie Sisley, Stavra Xanthakos, Rohit Kohli, Sarah Rock, Sandrah P. Eckel, Michele A. La Merrill, Max M. Aung, Matthew P. Salomon, Rob McConnell, Jesse Goodrich, David V. Conti, Lucy Golden-Mason, Lida Chatzi

## Abstract

To address the growing epidemic of liver disease, particularly in pediatric populations, it is crucial to identify modifiable risk factors for the development and progression of metabolic dysfunction-associated steatotic liver disease (MASLD). Per- and polyfluoroalkyl substances (PFAS) are persistent ubiquitous chemicals and have emerged as potential risk factors for liver damage. However, their impact on the etiology and severity of MASLD remains largely unexplored in humans. This study aims to bridge the gap between human and in vitro studies to understand how exposure to perfluoroheptanoic acid (PFHpA), one of the emerging PFAS replacements which accumulates in high concentrations in the liver, contributes to MASLD risk and progression. First, we showed that PFHpA plasma concentrations were significantly associated with increased risk of MASLD in obese adolescents. Further, we examined the impact of PFHpA on hepatic metabolism using 3D human liver spheroids and single-cell transcriptomics to identify major hepatic pathways affected by PFHpA. Next, we integrated the *in vivo* and *in vitro* multi-omics datasets with a novel statistical approach which identified signatures of proteins and metabolites associated with MASLD development triggered by PFHpA exposure. In addition to characterizing the contribution of PFHpA to MASLD progression, our study provides a novel strategy to identify individuals at high risk of PFHpA-induced MASLD and develop early intervention strategies. Notably, our analysis revealed that the proteomic signature exhibited a stronger correlation between both PFHpA exposure and MASLD risk compared to the metabolomic signature. While establishing a clear connection between PFHpA exposure and MASLD progression in humans, our study delved into the molecular mechanisms through which PFHpA disrupts liver metabolism. Our *in vitro* findings revealed that PFHpA primarily impacts lipid metabolism, leading to a notable increase of lipid accumulation in human hepatocytes after PFHpA exposure. Among the pathways involved in lipid metabolism in hepatocytes, regulation of lipid metabolism by PPAR-a showed a remarkable activation. Moreover, the translational research framework we developed by integrating human and in vitro data provided us biomarkers to identify individuals at a high risk of MASLD due to PFHpA exposure. Our framework can inform policies on PFAS-induced liver disease and identify potential targets for prevention and treatment strategies.

## Introduction

Metabolic dysfunction-associated steatotic liver disease (MASLD), previously known as nonalcoholic liver disease (NAFLD)^1^, refers to a spectrum of liver disorders including the metabolic dysfunction-associated steatohepatitis (MASH), formerly known as nonalcoholic steatohepatitis (NASH). A hallmark of MASLD is the fat accumulation (steatosis) in the liver due to chronic metabolic dysfunction^2^. MASLD management addresses long-term healthcare needs and generates substantial economic burden^3^. The prevalence of MASLD in children has been growing in recent years, paralleling the rise in childhood obesity and metabolic syndrome^4,5^. MASLD is now one of the most common chronic liver diseases in children worldwide, affecting approximately 10% of children in the general population^5^. Among children with overweight or obesity, the prevalence of MASLD is much higher, to an estimated of 30-40%^4^. Limited interventions are available for the improvement of MASLD in children ^6^. Diet restrictions, physical activity interventions and FDA-approved drugs including GLP-1 receptor agonists^7^ have been used with limited success in adolescents with MASLD^8^. This highlights the need for preventive measures, such as identifying and intervening on modifiable risk factors.

The traditional risk factors for MASLD, such as excess energy intake, sedentary lifestyle, and genetics, cannot fully explain the MASLD epidemic in children^9^. Moreover, emerging evidence indicates that exposure to endocrine-disrupting chemicals can promote metabolic changes that result in fatty liver disease - a hypothesis referred to as the ‘Toxicant Fatty Liver Disease’^10–12^. Per- and polyfluorinated substances (PFAS), a large class of synthetic fluorinated organic chemicals, are globally ubiquitous. These chemicals have been used in industrial applications and consumer products, including water-repellent textiles, nonstick coatings, and food packaging products, for over 60 years^13^. PFAS have been detected in blood of over 99% of individuals in the US.^14,15^ Production of certain PFAS, such as perfluorooctane sulfonate (PFOS) and perfluorooctanoate (PFOA), was voluntarily phased-out in the U.S. during the 2000s, yet their negative health effects remain a concern because of their long half-lives (1.8–6.2 years).^14,16–18^ Consequently, newer PFAS variants, known as replacements, have been introduced, featuring shorter biological half-lives, to mitigate environmental persistence.^16^ However, many of these replacements lack regulation and thorough testing regarding potential health risks, particularly during crucial developmental stages.

Additional studies are needed to better understand the health effects of replacement PFAS. Research has established that PFAS can accumulate in the human body and preferentially accumulate in the liver^19–21^, where they can disturb several hepatic functions, especially lipid metabolism^18,22–24^. An abundance of evidence exists based on concordance between experimental and population findings that certain PFAS are hepatotoxic to humans and many studies point to PFAS-induced lipid disruption^22^. Nevertheless, several gaps persist in the literature, such as a) whether individuals classified as overweight/obese are more prone to PFAS-induced liver toxicity, b) whether understudied PFAS compounds including replacement PFAS can induce liver damage, and c) which metabolic pathways affected by PFAS are significant and indicative of liver damage^18,22,24,25^.

We propose a translational research framework to connect scientific findings from human and in vitro studies, aiming to understand how PFAS contribute to MASLD progression (see Fig. 1). Our investigation focused on perfluoroheptanoic acid (PFHpA), a short-chain carboxylic PFAS compound found in high concentrations in the liver^23^ which was strongly linked to MASLD risk and severity of disease in obese adolescents. Using an innovative approach, we assessed PFHpA’s impact on liver metabolism in vitro by combining 3D human liver spheroids with single-cell transcriptomics. This allowed us to identify the primary metabolic pathways affected by PFHpA. Subsequently, we integrated multi-omic datasets from the human and in vitro study using advanced statistical methods. Through this analysis, we identified protein and metabolite signatures associated with MASLD development due to PFHpA exposure. Our study presents a novel strategy to identify individuals at high risk of developing PFAS-induced MASLD and to develop early intervention strategies.

**Figure 1.**
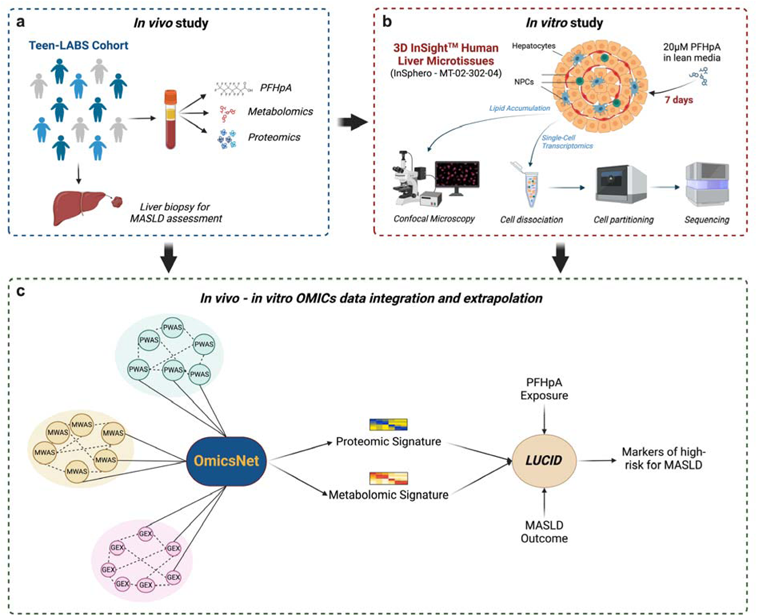
Translational framework. (a) Teen-LABS study design (b) Human liver spheroids composed of primary hepatocytes and non-parenchymal cells (NPCs) were exposed to 20µM PFHpA for 7 days. Culture media was changed every 2-3 days to ensure constant PFHpA presence in culture media. At the end of the culture period, spheroids were dissociated into single cells, and viable cells were partitioned using the 10x Genomics platform. Single-cell gene expression libraries were sequenced using the Illumina platform. Created with <u>BioRender.com.</u> (c) Data integration workflow. We used the OmicsNet 2.0 platform to integrate 156 differentially expressed genes upregulated in PFHpA-exposed spheroids (GEX), 42 metabolites (MWAS), and 28 proteins (PWAS) upregulated in the plasma of MAFLD subjects (compared to non-MAFLD controls). After analyzing the overlapped pathways between GEX-MWAS and GEX-PWAS, we identified 19 metabolites and 6 proteins that will be used for further analysis. Created with <u>BioRender.com</u>

## Results

### Human Study: PFHpA increases the risk for MASLD in adolescents – Insights into Steatosis Severity and Disease Progression

The current project included 136 adolescents with severe obesity who underwent bariatric surgery. Based on liver biopsies fifty-five (40%) participants were categorized as non-MASLD, 51 (38%) as MASLD not MASH, and 30 (22%) as MASH. Among 8 PFAS congeners, plasma-PFHpA (mean = 0.13 ng/mL, SD = 0.12 ng/mL) was the sole PFAS congener significantly associated with both MASLD and MASH (Fig. S1, Table S1). Among adolescents in the Teen-LABS study, we observed 68% higher odds of MASLD per doubling of PFHpA (OR: 1.68, 95% CI: 1.19, 2.37) (Fig. S1, Table S1). We observed a significant dose-dependent relationship with PFHpA (p for trend=0.002), indicating a biological gradient in MASLD risk across exposure octiles (Fig. 2b). Furthermore, PFHpA exhibited significant associations with several indicators of disease severity, including the degree of steatosis (OR=1.61, 95% CI: 1.16, 2.24 for mild steatosis and OR=1.88, 95% CI: 1.22, 2.89 for moderate steatosis), fibrosis (OR =1.49, 95% CI: 1.04, 2.14), and MASLD activity score (MAS) (OR=3.04, 95% CI: 1.78, 5.21). (Fig. 2c).

**Figure 2.**
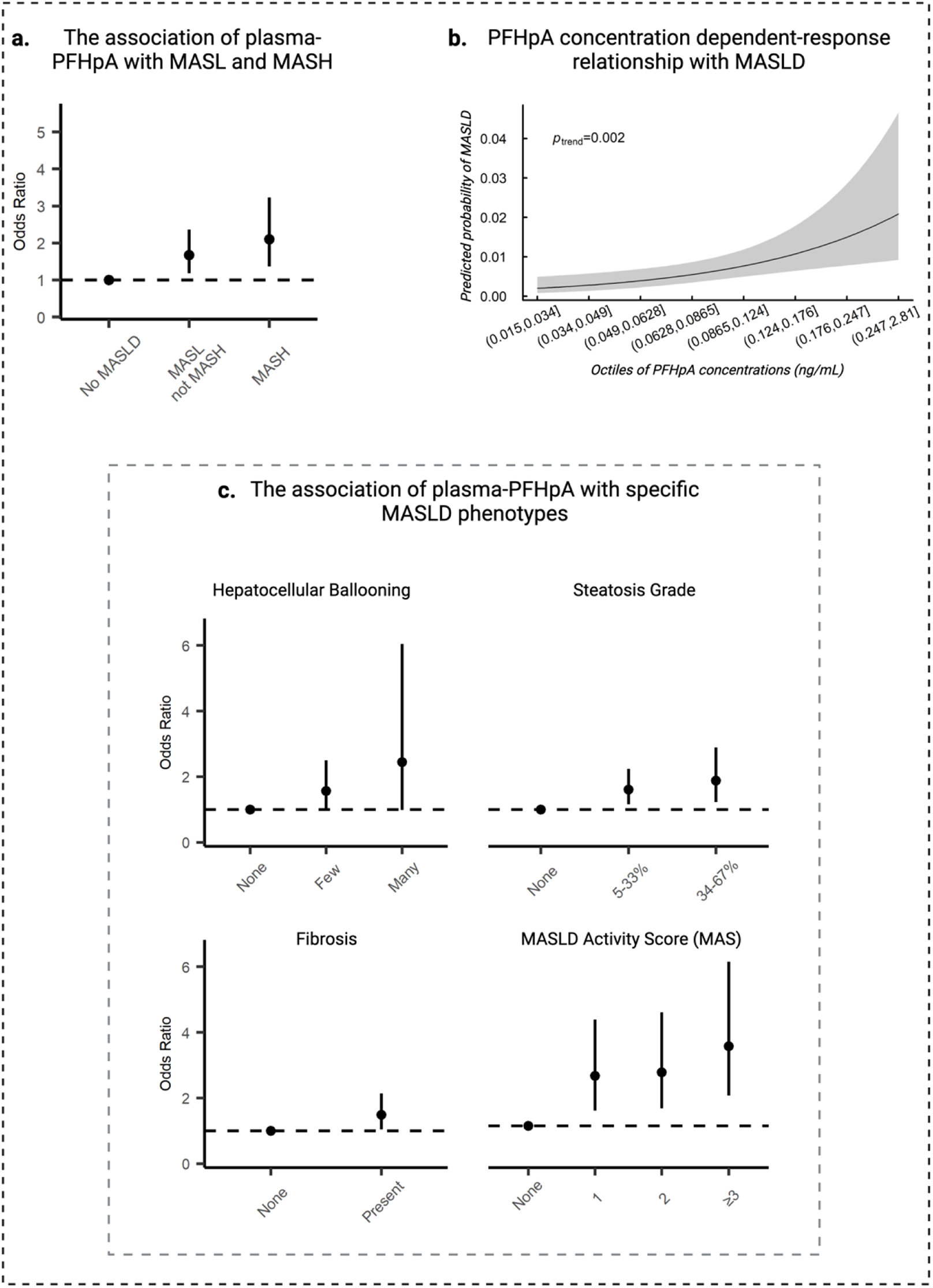
PFHpA exposure, histopathological determined outcomes of MASLD in the Teen-LABS study. (a) Shows the odds ratio (OR) and 95% confidence intervals (95% CIs) for the association between PFHpA (ng/mL) and histopathological determined MASLD (N=136). Multinomial logistic regression between PFHpA and severity of MASLD (No MASLD, MASL not MASH, MASH). The models controls for: age, sex, race, parental income, study site. Results are shown as log2 of PFHpA exposure and therefore interpreted as per doubling of PFHpA exposure. (b) The association between octiles of PFHpA and histopathological determined MASLD (N=136). Model adjusted for: age, sex, race, parental income, study site. Results are shown as log2 of PFHpA exposure and therefore interpreted as per doubling of PFHpA exposure. (c) The association between PFHpA and histopathological determined outcomes of SLD (N=136). Figure 1c shows the odds ratio (OR) and 95% confidence intervals (95% CIs) for the association between PFHpA (ng/mL) measured in plasma and refined outcomes of SLD. Multinomial logistic regression between PFHpA and hepatocellular ballooning, steatosis grade, and MASLD Activity Score (MAS); Logistic regression between PFHpA and presence (y/n) of fibrosis. All models adjusted for: age, sex, race, parental income, study site. Results are shown as log2 of PFHpA exposure and therefore interpreted as per doubling of PFHpA exposure.

### Human Study: PFHpA Affects Lipid Metabolism, Oxidative Stress, and Inflammation in Adolescents

Using Metabolome-Wide Association Study (MWAS) and Proteome-Wide Association Study (PWAS) approaches, we identified significant alterations in 51 metabolites and 55 proteins significantly altered by PFHpA exposure (Fig. 3a & 3b). Ingenuity Pathway Analysis (IPA) revealed the biological pathways affected by PFHpA (Fig. 3c & 3d). Specifically, pathways related to inflammation were enriched, including the Inflammation pathway influenced by altered metabolites, and the Inflammatory Response and Cell Movement of Dendritic Cells pathways affected by altered proteins. We also observed an increase in the Synthesis of Reactive Oxygen Species pathway by metabolites, underscoring the role of inflammation and oxidative stress in PFHpA toxicity. Additionally, disruptions in lipid metabolism were evident, with enrichment of pathways such as Synthesis of Lipid and Concentration of Lipid, indicating lipid dysregulation as a potential consequence of PFHpA exposure. PWAS IPA analysis further emphasized PFHpA’s associations with immune response, showing enrichment in immunity-related pathways by PFHpA-associated proteins. These pathways include Recruitment of Mononuclear Leukocytes, Formation of Lymphoid Tissue, Cell Proliferation of T Lymphocytes, and Cell Death of Lymphatic System Cells.

**Figure 3.**
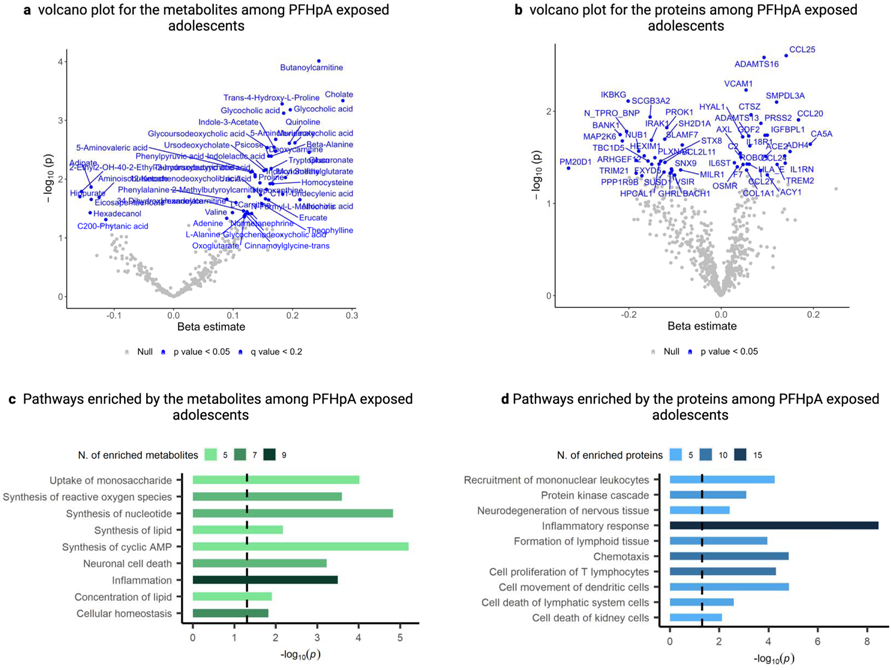
Omics-Wide Association Study in Teen-LABS. (a) Volcano plot showing the MWAS for PFHpA and metabolites from adolescents in Teen-LABS. Linear regression between PFHpA and metabolites (N=131) adjusting for age, sex, race, parental income, study site. (b) Volcano plot showing the PWAS for PFHpA and proteins (N=131) from adolescents in Teen-LABS. Linear regression between PFHpA and proteins (N=131) adjusting for age, sex, race, parental income, study site. (c) Enriched pathways from the MWAS for PFHpA and metabolites from adolescents in Teen-LABS. Linear regression between PFHpA and metabolites (N=131) adjusting for age, sex, race, parental income, study site. (d) Enriched pathways from the PWAS for PFHpA and proteins (N=131) from adolescents in Teen-LABS. Linear regression between PFHpA and proteins (N=131) adjusting for age, sex, race, parental income, study site.

### *In vitro* study: PFHpA disturbs lipid metabolism in human liver spheroids

To determine the involvement of PFHpA in MASLD progression, we performed an *in vitro* assay to expose human liver spheroids comprised of human primary hepatocytes and non-parenchymal cells to PFHpA for 7 days in low-glucose media (Fig. 1b). Subsequent analysis through single-cell RNA sequencing (scRNA-seq) revealed the transcriptomic alterations in the cells composing the liver spheroids. Of note, we identified hepatocytes, T cells, Kupffer cells and NK cells (Fig. 4a). B cells were detected in a very reduced number and therefore were not included in the further analysis. Overall, we found that PFHpA exposure altered the expression of 472 genes in liver spheroids, with 156 genes upregulated and 316 genes downregulated. Analyzing the hepatic cell populations, we observed that hepatocytes and T cells exhibited the most pronounced numbers of differentially expressed genes (DEGs), with 263 (137 up and 126 down) and 383 (98 up and 285 down) DEGs respectively (Fig. 4b). NK cells showed 26 DEGs (19 up and 7 down) and Kupffer cells only showed the upregulation of 1 gene. Pathway analysis using IPA highlighted a notable impact of PFHpA on liver metabolism. In the whole liver spheroids, as well as the hepatocytes and T cells, we observed a substantial upregulation of pathways involved in lipid metabolism (48%, 45%, and 40% respectively), amino acid metabolism (21%, 18%, and 20% respectively), and detoxification pathways (14%, 14%, and 10% respectively) indicating a significant disturbance induced by PFHpA exposure (Fig. 4c-e). Furthermore, PFHpA exposure led to the upregulation of peroxisome proliferator-activated receptor alpha (PPAR-α) activation and fatty acid oxidation in human hepatocytes (Fig. 4f). Most pathways involved in hepatocyte lipid metabolism were related to lipid anabolism (lipogenesis), with the upregulation of several lipid biosynthesis, including cholesterol. This observation was confirmed by Nile Red imaging of spheroid lipid accumulation (Fig. 4g), which showed that liver spheroids exposed to PFHpA significantly accumulate more lipids than control spheroids (p=0.01) (Fig. 4h). Although PFHpA exposure also activates lipid metabolism in T cells, the upregulated pathways were primarily related to nuclear hormone receptors activation, without a direct impact on lipid biosynthesis as observed in hepatocytes (Fig. S2). These findings underscore the pivotal role of PFHpA in disrupting lipid metabolism, particularly in hepatocytes, shedding light on the potential PFHpA contribution to MASLD pathogenesis.

**Figure 4.**
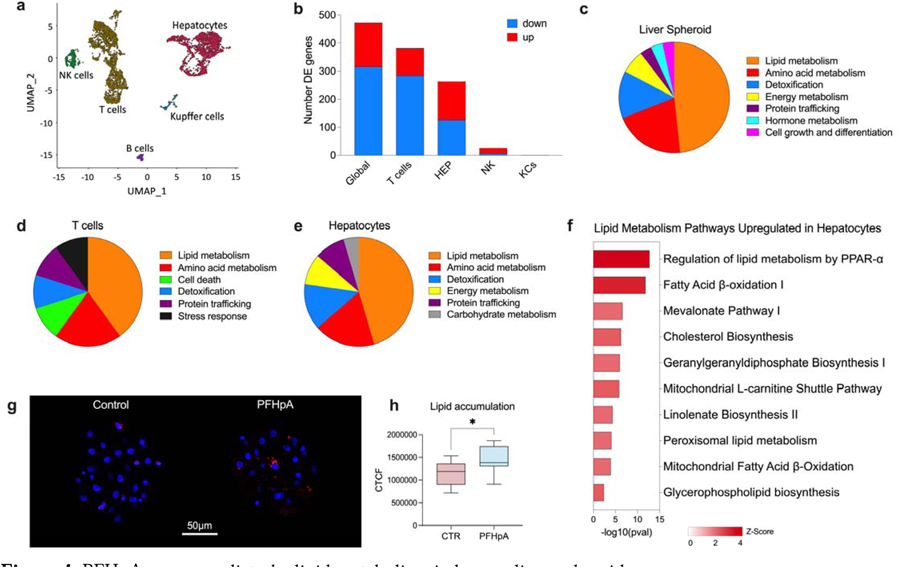
PFHpA exposure disturbs lipid metabolism in human liver spheroids. (a) Integrated UMAP of control and PFHpA-exposed spheroids. We observed the presence of hepatocytes, Kupffer cells, T cells, NK cells, endothelial cells, and B cells. (b) Differentially expressed (DE) genes detected in whole liver spheroid (global) and individual cell clusters. (c) Canonical pathways upregulated by PFHpA in whole liver spheroids. Around 48% of the pathways uploaded by PFHpA were related to lipid metabolism, while 20% were related to amino acid metabolism. Other important pathways were related to detoxification (13.8%), energy metabolism (6.9%), protein trafficking, hormone metabolism, and cell growth and differentiation. (d) Canonical pathways upregulated by PFHpA in T cells. Most of the pathways upregulated in T cells were related to lipid metabolism (40%) and amino acid metabolism (20%). Other pathways were related to cell death, detoxification, protein trafficking, and stress response. (e) Canonical pathways upregulated by PFHpA in hepatocytes. Almost 45% of the pathways upregulated in hepatocytes were related to lipid metabolism, and around 18% of the pathways were related to amino acid metabolism. Other important categories upregulated were energy metabolism, protein trafficking, and carbohydrate metabolism. (f) Upregulated pathways related to lipid metabolism in hepatocytes from spheroids exposed to PFHpA. (g) Lipid accumulation in liver spheroids. Confocal imaging of spheroids stained with Nile Red suggests an increase in lipid accumulation (red) in cells from liver spheroids exposed to PFHpA. (h) Digital quantification of confocal imaging using ImageJ confirms a significant increase in lipid accumulation due to PFHpA exposure. CTCF = Correlated Total Cell Fluorescence

### Unveiling Biomarker Signatures for PFHpA-induced MASLD through Multi-omics Integration

We integrated data from the human and the in vitro studies to identify plasma biomarkers indicative of PFHpA-induced MASLD using the OmicsNet 2.0 platform. Specifically, our analysis combined 156 upregulated genes (GEX) from the in vitro study with 42 metabolites (MWAS) and 28 proteins (PWAS) upregulated in the human study (Fig. 1c). OmicsNet 2.0 identified pathways showing significant overlap between GEX-MWAS and GEX-PWAS datasets. We identified a metabolomic signature of PFHpA-induced MASLD consisting of 19 metabolites involved in lipid metabolism, amino acid metabolism, nutrient uptake, energy metabolism, and vitamin and nucleic acid metabolism (Fig. 5b). Additionally, a proteomic signature was defined, comprising 6 proteins from pathways involved in lipid degradation, vitamin metabolism, immune response, detoxification, and cancer (Fig. 5c).

**Figure 5.**
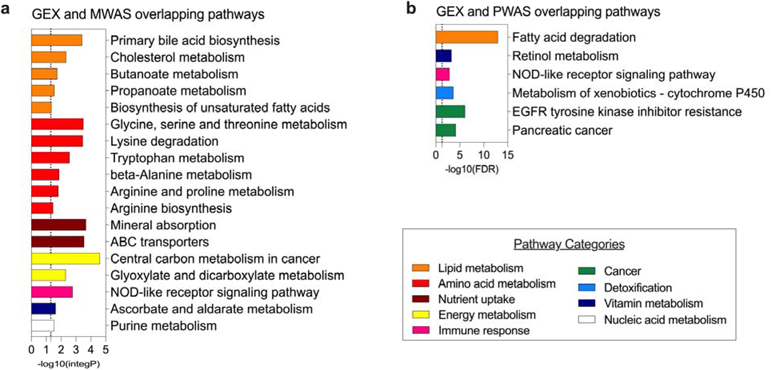
Integration of in vitro and in vivo datasets for identification of proteomic and metabolomic signatures. (a) Pathways commonly found in GEX (in vitro) and MWAS (in vivo) datasets. The shown pathways are composed of genes and metabolites. The majority of the pathways are related to lipid metabolism and amino acid metabolism. (b) Pathways commonly found in GEX (in vitro) and PWAS (in vivo) datasets. The shown pathways are composed of genes and proteins. The most significant pathway is related to lipid metabolism (fatty acid degradation [-log10(FDR)= 12.97].

Next, we integrated these overlapping features in a latent unknown clustering with integrated data (LUCID) model to assess the association between PFHpA exposure, multi-omics signatures, and MASLD risk. ^26^ This approach categorized individuals into groups based on similarities across PFHpA exposure, proteome, metabolome signatures, and disease outcomes, focusing on disease risk rather than stratification.

Figure 6 illustrates the joint associations between PFHpA and two latent clusters (profiles) of each omic layer (metabolome and proteome), identifying low and high MASLD risk clusters using unsupervised LUCID and logistic regression models . PFHpA showed a stronger association with the high-MASLD risk cluster characterized by proteins (OR = 2.73) compared to its association with the low-risk cluster characterized by metabolites (OR = 1.05). Notably, individuals in proteome profile 1 had significantly higher odds of MASLD (OR = 7.08) than those in proteome profile 0, while individuals in metabolome profile 1 had lower odds of MASLD (OR = 0.51) compared to those in metabolome profile 0. This designates metabolome profile 0 and proteome profile 1 as high-risk multi-omic profiles.

**Figure 6.**
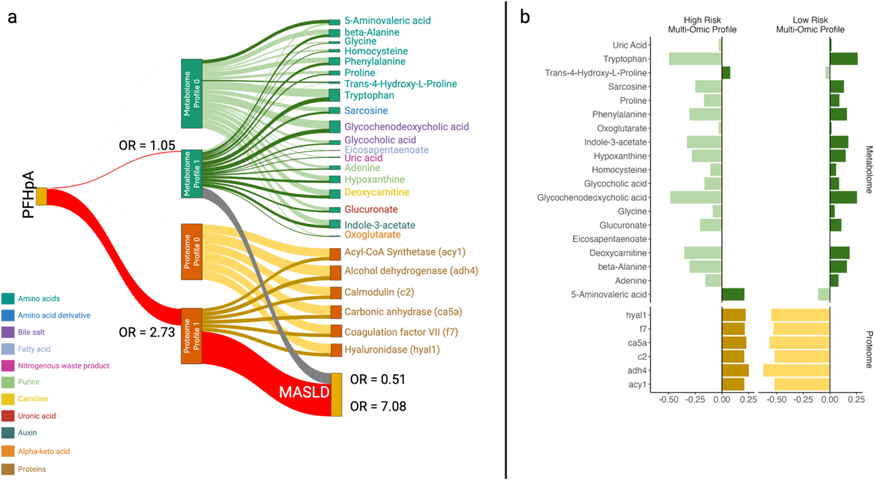
Multi-omics integration of PFHpA, proteomics, and metabolomics to determine clusters of individuals at high risk for MASLD (N=131). We identified two distinct multi-omic risk profiles associated with high PFHpA exposure and higher odds of MASLD. The first omic risk profile showed an association between high PFHpA levels and increased proteins. MASLD was 7 times more likely in this PFHpA-protein risk profile. The second omic risk profile showed an association between high PFHpA and altered levels of amino acids and lipids metabolites. (a) Shows the association of PIPS from unsupervised LUCID and MAFLD (No, Yes), N= 131. The model includes the two layers in the same model. REF = NO MASLD, model adjusted for study site, age, sex, race, parental income. The reference cluster for the proteins is the high-risk cluster and for the metabolites the reference cluster is the low-risk cluster. Results are shown as log2 of PFHpA exposure and therefore interpreted as per doubling of PFHpA exposure. (b) Demonstrates the high and low risk for MASLD groups of individuals by the clusters of metabolites and proteins. Results are shown as log2 of PFHpA exposure and therefore interpreted as per doubling of PFHpA exposure.

Fig. 6b describes the high and low risk for MASLD groups of individuals by the clusters of metabolites and proteins that differentiate these risk groups. Notably, tryptophan, glycochenodeoxycholic acid, and deoxycarnitine emerged as predominant metabolome features distinguishing high and low-risk profiles. Negative scaled values of metabolome features, with the exception of Trans-4-Hydroxy-L-Proline and 5-Aminovaleric acid, tended to correlate with a higher risk of MASLD. Conversely, positive scaled values of proteome features defined a higher risk of MASLD. Remarkably, HYAL1, F7, CA5A, C2, ADH4, and ACY1 exhibited similar scaled values ranging from 0.21 to 0.25 among high-risk profiles, while ranging from -0.63 to - 0.52 among low-risk profiles.

## Discussion

We demonstrated a significant link between exposure to PFHpA and the MASLD risk and severity of disease, and further elucidated the molecular and cellular mechanisms underlying this association. The Teen-LABS study is a highly unique resource—providing the opportunity to investigate the relationship between PFAS and MALD using a cohort with largely extreme obesity and consistent availability of well characterized liver biopsies for the diagnosis of MASLD^27^. Like other PFAS compounds, PFHpA is persistent in the environment and has been detected in water, air, soil, and biota worldwide. The half-life of PFHpA in humans is approximately 2 months^28^. PFHpA can be formed as a minor degradation product of long-chain PFAS^29–31^. Although PFHpA has gained attention in recent years, the relatively few studies investigating it may reflect a combination of factors including historical research priorities, technical challenges, and resource limitations. While previous research has established a connection between PFHpA exposure, along with other short and long-chain PFAS, and severity of liver steatosis and fibrosis in MASLD among adults^32^, our study marks the first time that exposure to PFHpA has been associated with MASLD in adolescents. Beyond the association with histological markers of MASLD, our findings indicate that adolescents with obesity exposed to PFHpA also exhibit alterations in plasma levels of proteins and metabolites that signal inflammation and dysfunction in lipid metabolism.

To delve deeper into the molecular mechanisms underlying PFHpA-associated MASLD, we conducted *in vitro* experiments to examine how PFHpA affects liver metabolism using the 3D human liver spheroid co-culture model. Our study used single-cell transcriptomics to evaluate PFHpA impact on each hepatic cell population from the liver spheroids. Our findings revealed that PFHpA primarily impacts lipid metabolism, leading to a notable increase in anabolic events in human primary hepatocytes, with significant hepatic lipid accumulation after PFHpA exposure. Among the pathways involved in lipid metabolism in hepatocytes, the ‘Regulation of lipid metabolism by PPAR-α’ and ‘Fatty Acid β-oxidation I’ pathways showed the strongest activation. This suggests that PFHpA promotes hepatocyte lipogenesis likely through peroxisome proliferator-activated receptor (PPAR)-α signaling. PPARs are members of the nuclear hormone receptor superfamily, acting as ligand-activated transcription factors^33^. In the liver, PPAR-α plays a crucial role in regulating fatty acid oxidation, and lipid and lipoprotein metabolism^34^, and previous research has shown that various PFAS, including PFHpA, can activate PPAR-α in cell lines and animal models^35–37^. Recently, Yang et al proposed that hepatic lipid metabolism disruption caused by PFOA and PFOS depends on the PPAR-α/ACOX1 axis^38^. Our *in vitro* analysis indicates this pathway is disrupted in hepatocytes but not immune cells. We found that ACOX1 expression was upregulated and integrated the pathway ‘Regulation of lipid metabolism by PPAR-α’ in hepatocytes from spheroids exposed to PFHpA. In contrast, while PPAR-α signaling was also upregulated in T cells exposed to PFHpA, ACOX1 was not differentially expressed and no lipid biosynthesis pathways were detected (Excel Supplement B). Our findings indicate that PFHpA alter signaling of PPAR-a/ACOX1 axis in hepatocytes but not T cells, to instigate abnormal hepatic lipid metabolism in humans.

Leveraging a novel statistical approach, we integrated *in vivo* and *in vitro* datasets to identify the proteomic and metabolomic signatures that could be used as potential indicators of PFHpA-induced MASLD. LUCID analysis was then applied to investigate whether these PFHpA-derived OMICs biomarkers correlated with disease outcome. Notably, our analysis revealed that the proteomic signature exhibited a stronger correlation between PFHpA exposure and MASLD risk compared to the metabolomic signature.

In addition to establishing a clear connection between PFHpA exposure and MASLD disease in humans, our study elucidated the intricate molecular mechanisms by which PFHpA impacts liver metabolism. Moreover, we developed a translational research framework that allows us to identify individuals at high risk of MASLD due to PFHpA exposure. Our framework not only advances the current knowledge on PFAS impact on chronic liver diseases but also provides critical information for future policies aiming at mitigating the detrimental impact of PFAS on human health. Finally, our findings offer potential new targets for MASLD treatment strategies by determining specific molecular targets implicated in PFAS-induced liver disease.

## Methods

### Study population

The study is based on data from the Teen-LABS study (ClinicalTrials.gov number, NCT00465829), a prospective, multicenter, observational study of adolescents (≤19 years of age) who underwent bariatric surgery from 2007 through 2012. The participants were enrolled at five clinical centers in the United States: Cincinnati Children’s Hospital Medical Center (Cincinnati, Ohio), Nationwide Children’s Hospital (Columbus, Ohio), the University of Pittsburgh Medical Center (Pittsburgh, Ohio), Texas Children’s Hospital (Houston, Texas) and the Children’s Hospital of Alabama (Birmingham, Alabama)^27^. Study inclusion criteria included (1) adolescents up to age 19, (2) adolescents approved for bariatric surgery, (3) agreement to participate in the Teen-LABS study, demonstrated through the signing of Informed Consent/Assent^27^. The Teen-LABS steering committee, which included a site principal investigator from each participating center, worked in collaboration with the data coordinating center and project scientists from the National Institute of Diabetes and Kidney Disease (NIH-NIDDK) to design and implement the study^27^. All bariatric procedures were performed by surgeons who were specifically trained for study data collection (Teen-LABS–certified surgeons)^27,39–41^. The present study includes 136 participants with plasma collected at the time of surgery. The study protocol, assent/consent forms, and data and safety monitoring plans were approved by the Institutional Review Boards of each institution and by the independent data and safety monitoring board prior to study initiation^27^. Written informed consent or assent, as appropriate for age, was obtained from all parents/guardians and adolescents^27^. Additionally, the study was approved by the University of Southern California review board.

### Data collection

Standardized methods for data collection have been described previously^27,39–41^. Fasting blood specimens were drawn at the preoperative visit. Liver histology and liver biopsy methodology has been previously detailed^41^, however, briefly, liver biopsies were obtained by core needle technique after induction of anesthesia and before performing the bariatric surgery procedure. Due to the observational study design and lack of published consensus on whether intra-operative liver biopsies should be standard of care at time of bariatric surgery, the decision to perform a liver biopsy was deferred to the surgical teams at each site. Accordingly, 99% of all biopsies were performed at the sites where intraoperative biopsy was standard of care. Liver biopsy specimens were stained with hematoxylin-eosin and Masson’s trichrome stains and reviewed and scored centrally by an experienced hepatopathologist using the validated MASH Clinical Research Network scoring system^42^. MASLD was defined by the histopathological diagnosis. Detailed descriptions of study methods, comorbidity and other data definitions, case report forms, and laboratory testing are included in previous publications^27,39,40^. For this analysis, covariates, including participants’ age, sex assigned at birth, race, and parents’ income, were obtained at the time of surgery with trained study personnel^27,39^. Collected data were maintained in a central database by the data coordinating center.

### Plasma-PFAS Laboratory Analysis

The samples were transported on dry ice with temperature logging by World Courier (AmerisourceBergen Corporation, Conshohocken, PA), and stored at - 80°C until analysis. The samples were analyzed by on-line solid phase extraction followed by LC-MS/MS as previously described^43,44^. The LOD was 0.03 ng/mL for all the reported compounds. Values below the LOD were imputed as ½ LOD. The batch imprecision for the quality controls was better than 6%.

### Plasma Metabolomics

Untargeted plasma metabolomics were measured in plasma samples collected at the time of bariatric surgery. Liquid chromatography coupled with high-resolution mass spectrometry methods (LC-HRMS) was used as described in Liu, et al.,^45^ with dual column and dual polarity approaches and both positive and negative electrospray ionization. This resulted in four analytical configurations: reverse phase (C18) positive, C18 negative, hydrophilic interaction (HILIC) positive, and HILIC negative. Unique features were identified using mass-to-charge ratio (m/z), retention time, and peak intensity. Features were adjusted for batch variation^46^ and excluded if they were detected in < 20% of samples or if there was a > 30% coefficient of variability of the quality control samples after batch correction. After processing, there were 3,716 features from the C18 negative mode, 5,069 from the C18 positive mode, 7,444 from the HILIC negative mode, and 6,944 from the HILIC positive mode, for a total of 23,173 features included in the analyses. The raw intensity values from LC-HRMS were scaled to a standard normal distribution and log_2_ transformed. Details of the analytical process have been described previously^47^. Confirmed annotations with confidence level 1 were available for 358 metabolomic features^48^. Metabolites were identified by comparing them to authentic chemical standards under identical analytical conditions, and peaks were matched to annotations using m/z and retention time. In instances where multiple annotations were possible due to more than one molecule having retention times within the allowable error, the annotation with the closest retention time to the known standard was chosen. Measured m/z and retention times, theoretical m/z and retention times, adduct, possible annotations, and additional analytical details are listed in Excel Supplement A.

### Plasma Proteomics

Proteins were measured in fasting plasma samples using the proximity extension array (PEA) method from Olink Explore 384 Cardiometabolic panel and Olink Explore 384 Inflammation panel^49^. These panels measure the relative abundance of 731 proteins, reported as normalized protein expression (NPX) levels after log_2_ transformation^50^. After excluding proteins with over 50% of observations below the limit of detection (LOD) and duplicate proteins, we retained 702 proteins from the initial 731 offered after processing.

### Liver spheroid assay

We used the 3D InSight™ Human Liver Model (MT-02-302-04, InSphero Inc.) to test the impact of PFHpA on human liver metabolism. This model is composed of human primary hepatocytes from 10 donors (5 male and 5 female), and non-parenchymal cells from 1 donor. PFHpA (CAS#375-85-9, Sigma-Aldrich, cat# 342041) stock solution was prepared in dimethylsulfoxide (DMSO, Sigma-Aldrich). The final working solution was diluted in lean spheroid media (CS-07-305B-01, InSphero Inc.) to a final non-cytotoxic concentration of 20µM of PFHpA and 0.1% DMSO^51^. Liver spheroids were continuously exposed to PFHpA for 7 days, and culture media was replaced every 2-3 days by media containing fresh diluted PFHpA. For control, liver spheroids were exposed to 0.1% DMSO diluted in lean spheroid media cultured for 7 days following the same regimen of media replacement. Spheroids were cultured in a 96-well format, with a single spheroid per well, under sterile conditions and incubated at 37°C, 5% CO_2_ following instructions provided by the manufacturer. We used 96 spheroids per condition.

### Lipid accumulation assay and analysis

After 7 days of treatment, spheroids were fixed with 4% paraformaldehyde in phosphate-buffered saline (PBS) for 1h, permeabilized with 0.2% Triton-X100 in PBS for 30 min and blocked with 1% bovine serum albumin (BSA) in PBS for 1h at room temperature. Then, spheroids were stained with Nile Red (1µg/mL) and DAPI (2µg/mL) in 1% BSA/PBS for 30 min and washed 3x with PBS. Spheroids were mounted with Prolong Gold Mounting Medium (Invitrogen) and images were acquired using a Leica TCS SP8 confocal microscope. Lipid accumulation quantification was performed using ImageJ^52^.

### Single-cell RNA library preparation, sequencing, and data analysis

Spheroids were dissociated using 0.25% trypsin for 15 min and dead cells were removed by Dead Cell Removal Kit (Miltenyi Biotech). Viable cells were partitioned with Chromium Next GEM Single Cell 3 Kit (10X Genomics). Libraries were sequenced at the USC Molecular Genomics Core using the Illumina platform. Raw data was processed using the Cell Ranger count pipeline (10X Genomics), with low-quality cell removal according to sample-specific quality control (QC) using the R package Seurat^53^. Comparison between PFHpA-treated and control samples was performed by integrating samples into a unified data set using the SCTransform integration workflow implemented in Seurat^54^. Cell annotation was based on the expression of known cell type marker genes. Differentially expressed genes between clusters from treatment groups were detected using the Wilcoxon Rank Sum test in the Seurat FindMarkers and genes with a Bonferroni adjusted p-value < 0.1 were considered significant. Biological data interpretation was performed using Ingenuity Pathways Analysis (IPA)^55^.

### Multi-omics data integration

We performed knowledge-driven integration of the differentially expressed genes obtained from transcriptomic PFHpA/liver spheroids in vitro assay (GEX), and the plasma proteomics (PWAS) and metabolomics (MWAS) data obtained from the Teen-LABS cohort using the online platform OmicsNet 2.0 (www.omicsnet.ca)^56^. Briefly, OmicsNet identified the significant canonical pathways in each dataset and further separately overlapped the results from GEX and PWAS, and GEX and MWAS. We then identified the proteins and metabolites part of the overlapped pathways and used them as Omics signatures for further analysis using LUCID.

## Statistical Analysis

### Plasma-PFHpA and MASLD

Plasma concentrations of PFAS measured in Teen-LABS participants has been previously published^23^. We first evaluated the associations of 8 plasma-PFAS with MASLD (yes/no) using logistic regression and controlling for multiple comparisons via a Bonferroni correction. The plasma-PFAS concentrations were log2 transformed. PFHpA was the only congener found to be associated with MASLD in our study; we therefore focused subsequent analyses on further exploring the association between plasma-PFHpA and MASLD and its features using either multinomial logistic regression models or logistic regression models, based on the number of categories in each outcome. The outcomes of interest included progression of MASLD (No MASLD, MASL not MASH, MASH; multinomial regression), hepatocellular ballooning (none, few, many; multinomial logistic regression), grade of steatosis (none, 5-33%, 34-67%; multinomial logistic regression), fibrosis (none, present; logistic regression), and MAS activity score (none, 1, 2, ≥3; multinomial logistic regression). For all models, we adjusted for participants’ age, race, sex assigned at birth, parents’ annual income, and site of medical center. To test the concentration dependent-relationship between plasma-PFHpA and MASLD, we performed a generalized additive model with a low rank thin plate spline. The continuous PFHpA concentration was categorized by octiles to mitigate the influence of extremely high exposures while maintaining a readily interpretable dose-response relationship.

### Metabolome-wide association study (MWAS)—linking PFHpA exposure with disease pathways

To perform the metabolome-wide analysis study, we included confirmed annotations with confidence level 1 metabolomic features^48^. The log-transformed metabolite intensity as included as the dependent variables and the log-transformed PFHpA concentration as the independent variable in a multiple linear regression model. We adjusted for participants’ age, race, sex assigned at birth, parents’ annual income, and site of medical centers to control for potential confounding. As we are not primarily interested in strict metabolite identification, we conducted an over-representation analysis for metabolites that were altered by PFHpA exposure (nominal *p* < 0.05) using canonical pathways and diseases and biofunctions curated from Qiagen Knowledge Base using QIAGEN Ingenuity Pathway Analysis (IPA, Qiagen Inc.). For IPA results (Fig. 3), we included the pathways with the number of enriched metabolites larger than or equal to five and kept the pathway with the lowest *p*-value in each category. As a result, nine pathways remained and were reported in the current analysis. Then, we used a liberal threshold of nominal *p* < 0.2 to select the metabolites entering the downstream OmicsNet step in order to include all possible putative metabolites that were involved in the underlying mechanisms.

### Proteome-wide association study (PWAS)—linking PFHpA exposure with disease pathways

To perform the proteome-wide analysis study, we included the NPX levels as dependent variables and the log-transformed PFHpA concentration as the independent variable in a multiple linear regression model. We adjusted for participants’ age, race, sex assigned at birth, parents’ annual income, and site of medical centers to control for potential confounding. For proteins that were differentially expressed by PFHpA exposure (nominal *p* < 0.05), we conducted the over-representation analysis by QIAGEN Ingenuity Pathway Analysis (IPA, Qiagen Inc.) for canonical pathways and diseases and biofunctions curated from Qiagen Knowledge Base. For IPA results (Fig. 3), we included the pathways with the number of enriched metabolites larger than or equal to five and kept the pathway with the lowest *p*-value in each category. In addition, as IPA yielded more pathways for PWAS than MWAS, we also used the absolute value of Z-score > 2 as a cutoff. As a result, ten top pathways with the lowest *p*-values were reported in the current analysis. Then, we used a threshold of nominal *p* < 0.2 to select the proteins entering the downstream OmicsNet step in order to include all possible proteins that were involved in the underlying mechanisms.

### Multi-omics integration-Latent Unknown Clustering by Integrating multi-omics Data (LUCID)

Latent Unknown Clustering by Integrating multi-omics Data (LUCID) is a novel quasi-mediation analysis approach of multi-omics data that estimates the joint associations between the environmental exposure E (PFHpA), the multi-omics data Z (19 metabolites and 6 proteins that were identified through prior pre-screening procedures), and the outcome Y (MASLD) if supervised via the latent cluster variable X. The Expectation-Maximization (EM) algorithm is implemented to iteratively estimate X and update the parameters until convergence. For an unsupervised LUCID model, parameters of interest include (1) f3, representing PFHpA-to-cluster associations; (2) µ, representing the cluster-specific means of omics features; and (3), the individual level Inclusion probability (IP) to each latent cluster. In the unsupervised LUCID approach, X integrates information from both E and Z, effectively delineating distinct risk profiles among subjects. The original LUCID framework was initially proposed for early integration of multi-omics data, entailing the concatenation of all omics layers into a single matrix. Detailed descriptions of the original LUCID have been previously introduced^26^. As an extension of the original LUCID, LUCID in parallel utilizes a intermediate integration strategy of the multi-omics data to estimate separate latent clusters X within each omic layer by assuming no correlations across different omics layers^57^. In our study, there were 2 layers of multi-omics data Z, metabolites and proteins, resulting in two individually estimated latent cluster variables, X_metabolome_ and X_proteome_. We fitted the optimal unsupervised LUCID in parallel model using PFHpA as E and pre-selected 19 metabolites and 6 proteins as Z. Based on model selection procedures using Bayesian information criterion (BIC), the number of latent clusters per omic layer of the optimal model was chosen to be 2. We extracted the IPs to X_metabolome_ and X_proteome_ (/P_metabolome_ and /P_proteome_) from the converged optimal LUCID in parallel model.

/P_metabolome_ and /P_proteome_ are continuous probabilities indicating the likelihood of being included in each level of cluster X_metabolome_ and X_proteome_, respectively. These probabilities are determined by the subjects’ exposure levels, as well as the presence of metabolites and proteins, respectively. In follow up analyses to explore how /P_metabolome_ and /P_proteome_ were associated with the outcome of interest, MASLD, we fitted a logistic regression model using /P_metabolome_ and /P_proteome_ as the predictors and MASLD as the binary response variable, while adjust for covariates including participants’ age, race, sex assigned at birth, parents’ annual income, and site of medical centers.

## Human Subjects

Ethics approval for this study was provided by the University of Southern California Institutional Review Board (IRB protocols HS-19-00057). Prior to participation, written informed assent/consent were obtained from participants and their guardians. The Teen–Longitudinal Assessment of Bariatric Surgery (Teen–LABS) study (<u>ClinicalTrials.gov</u> number, <u>NCT00474318</u>) was designed as prospective, multicenter, observational study of consecutive cases of bariatric surgery offered to adolescents. The study methodology has been previously described.

## Data Availability

Raw and processed scRNA-seq datasets were deposited into the NCBI GEO database under the accession number GSE253186. All Teen-LABS data and metadata are uploaded, stored and make available through the existing NIH-NIDDK Central Repository.

## Acknowledgements

The authors would like to acknowledge the significant contributions made by all Teen-LABS study personnel as well as study participants. Additionally, the authors would like to acknowledge the contributions made by Flemming Nielsen, PhD and Philippe Grandjean, MD, PhD—both lead the laboratory analysis of plasma-PFAS of Teen-LABS participants.

---

**Table 1.** Characteristics of Teen-LABS cohort (N=136)

|  |  |
| --- | --- |
| Age (years), mean (SD) | 17.1 (1.5) |
| White, n (%) | 93 (68.4%) |
| Female, n (%) | 100 (73.5%) |
| Parent's income, less than \$75,000, mean (SD) | 111 (81.6%) |
| BMI (kg/m<sup>2</sup>), mean (SD) | 53.8 (9.8) |
| BMI: Body Mass Index |  |

**Table 2.** Refined outcomes of SLD in Teen-LABS (N=136)

|  | n (%) |
| --- | --- |
| MASLD |  |
| No MASLD | 55 (40.4) |
| MASLD | 81 (59.6%) |
| MASLD severity |  |
| No MASLD | 55 (40.4) |
| MASLD, not MASH | 51 (37.5) |
| MASH | 30 (22.1) |
| MAFLD Activity Score (MAS) |  |
| none | 25 (18.4) |
| 1 | 41 (30.1) |
| 2 | 37 (27.2) |
| ≥3 | 33 (24.3) |
| Fibrosis |  |
| None | 110 (80.9) |
| Present | 26 (19.1) |
| Steatosis |  |
| None | 106 (77.9) |
| 5 - 33% | 18 (13.2) |
| 34 - >67% | 12 (8.9) |
| Hepatocellular ballooning |  |
| None | 115 (84.6) |
| Few | 16 (11.8) |
| Many | 5 (3.6) |

Metabolic dysfunction-associated steatotic liver disease (MASLD); Steatotic liver disease (SLD); The MAS can range from 0 to 8 and is calculated by the sum of scores of steatosis (0-3), lobular inflammation (0-3) and hepatocellular ballooning (0-2).

