## Supplemental Tables and Figures for "PFHpA alters lipid metabolism and increases the risk of metabolic dysfunction-associated steatotic liver disease in youth—a translational research framework"

Table of Contents

**Figure S1.** Association between plasma-PFAS and MASLD in Teen-LABS

**Table S1.** Association between plasma-PFAS and MASLD in Teen-LABS

**Table S2.** Metabolome-Wide Association Study (MWAS) between plasma-PFHpA and metabolites in Teen-LABS

**Table S3.** Proteome-Wide Association Study (PWAS) between plasma-PFHpA and proteins in Teen-LABS

**Figure S2.** Lipid Metabolism Pathways Upregulated in T cells

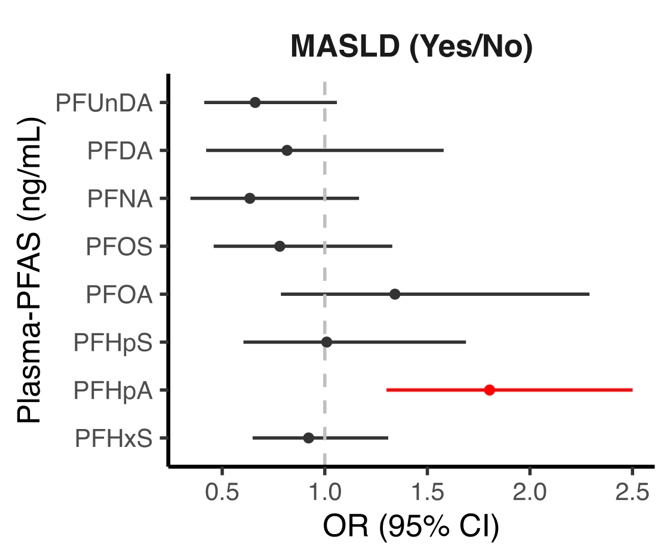

**Figure S1.** Association between plasma-PFAS and MASLD in Teen-LABS controlling

for race, sex, parent’s income, clinical site

| **Table S1.**  Association between plasma-PFAS and MASLD in Teen-LABS | | | | |
| --- | --- | --- | --- | --- |
|  | OR | 95% CI | | P Value |
| PFUnDA | 0.66 | 0.41 | 1.06 | 0.34 |
| PFDA | 0.82 | 0.42 | 1.58 | 0.73 |
| PFNA | 0.63 | 0.34 | 1.17 | 0.38 |
| PFOS | 0.78 | 0.46 | 1.33 | 0.58 |
| PFOA | 1.34 | 0.79 | 2.29 | 0.56 |
| PFHpS | 1.01 | 0.60 | 1.69 | 0.97 |
| PFHpA | 1.80 | 1.30 | 2.50 | 0.003 |
| PFHxS | 0.92 | 0.65 | 1.31 | 0.74 |
| Logistic regression controlling for race, age, sex, parent's income, clinical site | | | | |

| **Table S2**. Metabolome-Wide Association Study (MWAS) between plasma-PFHpA and metabolites in Teen-LABS | | | | |
| --- | --- | --- | --- | --- |
| **Metabolite name** | **estimate** | **95% CI** | | **p value** |
| Proline | 0.15 | 0.05 | 0.25 | 0.00 |
| 5-Aminovaleric acid | 0.07 | -0.02 | 0.15 | 0.11 |
| Glycine | 0.06 | -0.04 | 0.16 | 0.22 |
| Sarcosine | 0.09 | -0.01 | 0.19 | 0.07 |
| Trans-4-Hydroxy-L-Proline | 0.14 | 0.04 | 0.24 | 0.00 |
| Homocysteine | 0.14 | 0.02 | 0.25 | 0.02 |
| Hypoxanthine | 0.13 | 0.02 | 0.24 | 0.02 |
| Oxoglutarate | 0.10 | -0.01 | 0.21 | 0.06 |
| Deoxycarnitine | 0.09 | -0.02 | 0.19 | 0.10 |
| Phenylalanine | 0.05 | -0.07 | 0.18 | 0.41 |
| Uric Acid | 0.12 | 0.04 | 0.21 | 0.00 |
| Indole-3-acetate | 0.12 | 0.03 | 0.22 | 0.01 |
| Adenine | 0.07 | -0.03 | 0.17 | 0.19 |
| Glucuronate | 0.18 | 0.04 | 0.32 | 0.01 |
| Tryptophan | 0.09 | -0.02 | 0.19 | 0.12 |
| Eicosapentaenoate | -0.11 | -0.21 | -0.02 | 0.02 |
| Glycochenodeoxycholic acid | 0.13 | 0.03 | 0.23 | 0.01 |
| Glycocholic acid | 0.16 | 0.06 | 0.26 | 0.00 |
| beta-Alanine | 0.14 | 0.05 | 0.24 | 0.00 |
| MWAS between plasma-PFHpA and metabolites measured in Teen-LABS participants controlling for race, age, sex, parent's income, clinical site. A p-value threshold of 0.2 was used for inclusion in the multi-omics integration (OmicsNet) step of the framework. | | | | |

| **Table S3.** Proteome-Wide Association Study (PWAS) between plasma-PFHpA and metabolites in Teen-LABS | | | | |
| --- | --- | --- | --- | --- |
| **Protein name** | **estimate** | **95% CI** | | **p value** |
| acy1 | 0.10 | 0.01 | 0.18 | 0.02 |
| adh4 | 0.16 | 0.04 | 0.27 | 0.01 |
| c2 | 0.02 | -0.01 | 0.06 | 0.23 |
| ca5a | 0.19 | 0.05 | 0.33 | 0.01 |
| f7 | 0.03 | 0.00 | 0.07 | 0.08 |
| hyal1 | 0.03 | 0.00 | 0.07 | 0.05 |
| PWAS between plasma-PFHpA and proteins measured in Teen-LABS participants controlling for race, age, sex, parent's income, clinical site. A p-value threshold of 0.2 was used for inclusion in the multi-omics integration (OmicsNet) step of the framework. | | | | |

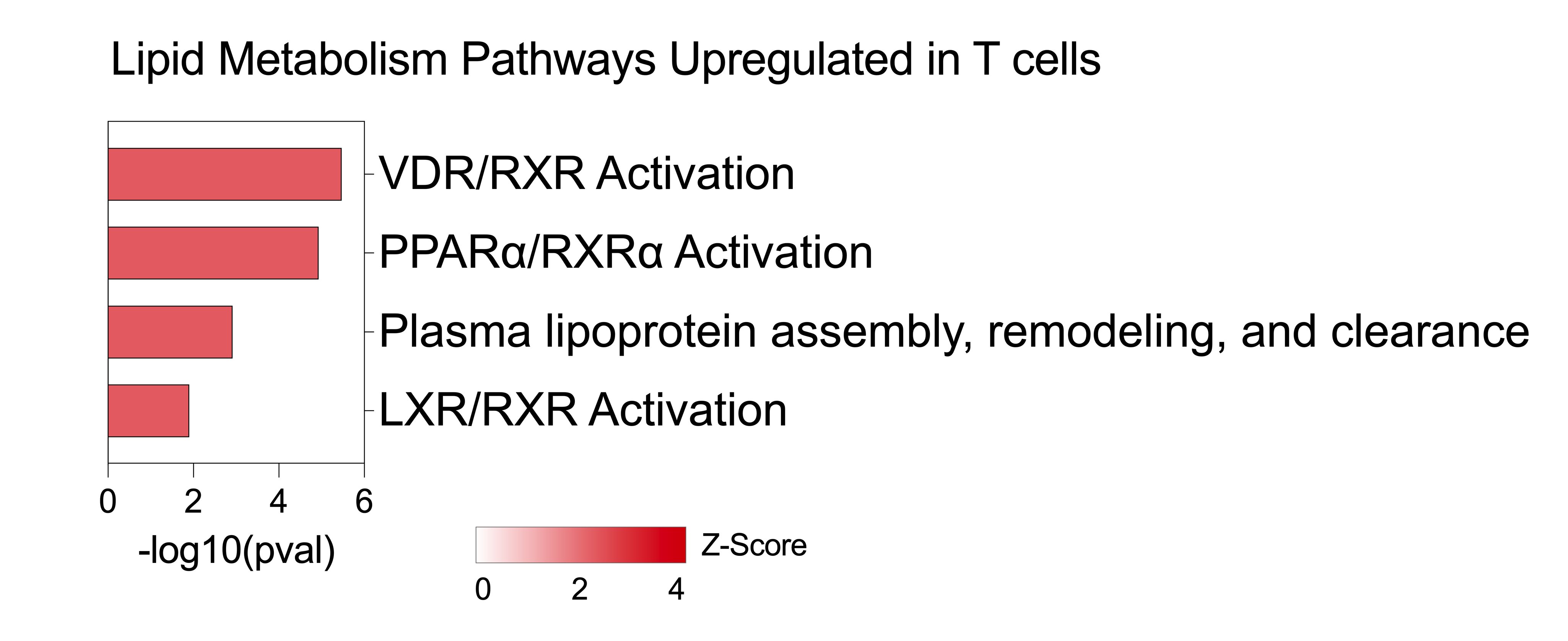

**Figure S2.** Lipid metabolism pathways upregulated in T cells from 3D liver spheroids exposed to PFHpA for 7 days.
